# Stroke recovery-related changes in cortical reactivity based on modulation of intracortical inhibition

**DOI:** 10.1101/2022.09.20.22280144

**Authors:** Sylvain Harquel, Andéol Cadic-Melchior, Takuya Morishita, Lisa Fleury, Adrien Witon, Martino Ceroni, Julia Brügger, Nathalie H. Meyer, Giorgia G. Evangelista, Philip Egger, Elena Beanato, Pauline Menoud, Dimitri Van de Ville, Silvestro Micera, Olaf Blanke, Bertrand Léger, Jan Adolphsen, Caroline Jagella, Christophe Constantin, Vincent Alvarez, Philippe Vuadens, Jean-Luc Turlan, Andreas Mühl, Diego San Millán, Christophe Bonvin, Philipp J. Koch, Maximilian J. Wessel, Friedhelm C. Hummel

## Abstract

**Objective:** Cortical excitation/inhibition dynamics have been suggested as a key mechanism occurring after stroke. Their supportive or maladaptive role in the course of recovery is still not completely understood. Here, we used TMS-EEG coupling to study cortical reactivity and intracortical GABAergic inhibition, as well as their relationship to residual motor function and recovery longitudinally in a large cohort of stroke patients.

**Methods:** EEG responses evoked by TMS applied to the ipsilesional motor cortex were acquired in 66 stroke patients in the acute (1-week), subacute (3-weeks) and early chronic (3-months) stage. Readouts of cortical reactivity and intracortical inhibition were drawn from TMS-evoked potentials induced by single pulse and paired pulse TMS. Residual function of the upper limb was quantified through a detailed motor evaluation.

**Results:** Most affected patients exhibited larger and simpler brain reactivity patterns. Bayesian statistics revealed a link between abnormally high motor cortical excitability in the acute stage and impairment level, while a decrease of excitability in the following months was related to better motor recovery. The investigation of the intracortical GABAergic inhibitory system using paired pulse TMS revealed the presence of a beneficial disinhibition in the acute stage, followed by a normalization of inhibitory activity.

**Interpretation:** The present results revealed an abnormal motor cortical reactivity in stroke patients, which was driven by perturbations and longitudinal changes within the intracortical inhibition system. They support the view that disinhibition in the ipsilesional motor cortex during the first week post-stroke is beneficial and promotes neuronal plasticity and recovery.

## Introduction

Although more knowledge is continuously gained on the neurobiological processes occurring in the first weeks and months after a stroke, the mechanisms sustaining motor improvement are still not fully understood^1^. There is substantial evidence that stroke induces functional plasticity partly driven by alterations in neuronal excitability^2–4^. Indeed, in the first phase after a stroke, strong release of glutamate is excitotoxic and contributes to cell death, which is counteracted by the inhibitory neurotransmitter GABA through cell hyperpolarization^5^. In mice, this phase in which inhibition in the perilesional area is beneficial lasts approximately 3 days^6^, while its duration in humans remains unknown^4^. In the longer term, the effects are eventually reversed, so that a shift in the cortical excitatory/inhibitory balance towards excitation becomes beneficial for plasticity^7,8^. The resulting increase in excitability has been associated to the induction of structural plasticity^9–13^ and functional reorganization in motor regions^14–16^.

Collectively, this evidence suggests that changes in this balance could be one pivotal mechanism at the origin of neural plasticity^4,17^. However, these mechanisms still need to be validated in longitudinal investigations *in vivo* in humans, to better understand the factors sustaining stroke recovery and to unveil potential targets for therapy tailored to the specific phase of the recovery process. Combining Transcranial Magnetic Stimulation (TMS) and scalp Electro-Encephalography (EEG) offers the possibility to directly assess the neuronal properties of the lesioned motor regions by studying the amplitude and dynamics of the TMS-Evoked Potentials (TEPs)^18^. Such properties include cortical excitability^19,20^, local cytoarchitectonics and remote connectivity^21–23^, and neurotransmitters concentrations such as GABA^24–26^.

TMS-EEG has been successfully applied in stroke. Ipsilesional motor cortical excitability was reported higher in stroke than in controls^27^ and was related to poorer motor function^28^. Moreover, an abnormal brain reactivity to TMS, defined by a large and simple monophasic evoked activity, was observed in the most affected patients^29,30^. This activity showed a similar profile as responses evoked in sleep and unresponsive wakefulness syndrome patients^31^. In addition, GABA receptors were suggested to be the main actors involved in this atypical brain reactivity. Crucially, TMS-EEG coupling also offers the benefit to directly investigate inhibitory mechanisms (GABA-ergic) by applying paired-pulse short-interval intracortical inhibition (SICI) TMS protocols^17,26,32–36^.

Here, we longitudinally evaluated (acute to early chronic) a cohort of stroke patients with TMS-EEG, in the framework of the TiMeS project^37^. The present study specifically focused on the analysis of the TMS-*evoked* responses and complements the study of TMS-*induced* oscillations^38^, the results of which will be put into perspective with those of the present study. Complementary TMS-EEG readouts allowed to study cortical excitability and evoked dynamics from the individuals’ brain reactivity, and their association with motor function at each stage and during the process of motor recovery. Additionally, by using for the first time SICI protocols in TMS-EEG coupling in stroke patients, we monitored the changes in intracortical inhibitory activity, and their relationship with residual motor function, impairment and recovery.

## Methods

### Patient population

Seventy-six stroke patients were enrolled in the study after being admitted at the cantonal hospital in Sion, Switzerland. Patients were recruited during the first week post-stroke, inclusion criteria consisted in being older than 18 y.o., motor deficits of the upper limb and absence of contraindications for magnetic resonance imaging (MRI) or TMS. Patients with first-ever or recurrent ischemic or hemorrhagic strokes were included. Among them, 66 patients (age: 68.2 ± 13.2 y.o., 18 females) were included in this study, i.e., patients with TMS-EEG recordings at least in one of the recording sessions (see Table 1 and Fig. 1.A for patients’ characteristics). With the aim of determining factors specific to recovery, the subset including patients showing motor improvement was defined as the recovering group (N = 40 in total). The latter was quantified by an increase in the Fugl-Meyer of the upper limb (see *Behavioral assessment* section below), from the acute to either of the following stages. 15 healthy older adults (aged-matched with patients, age: 67.0 ± 4.9 years, 11 females) were additionally recruited and underwent a single TMS-EEG recording session. The study was conducted in accordance with the Declaration of Helsinki and approved by Cantonal Ethics Committee Vaud, Switzerland (2018-01355), written informed consent was obtained.

**Figure 1.**
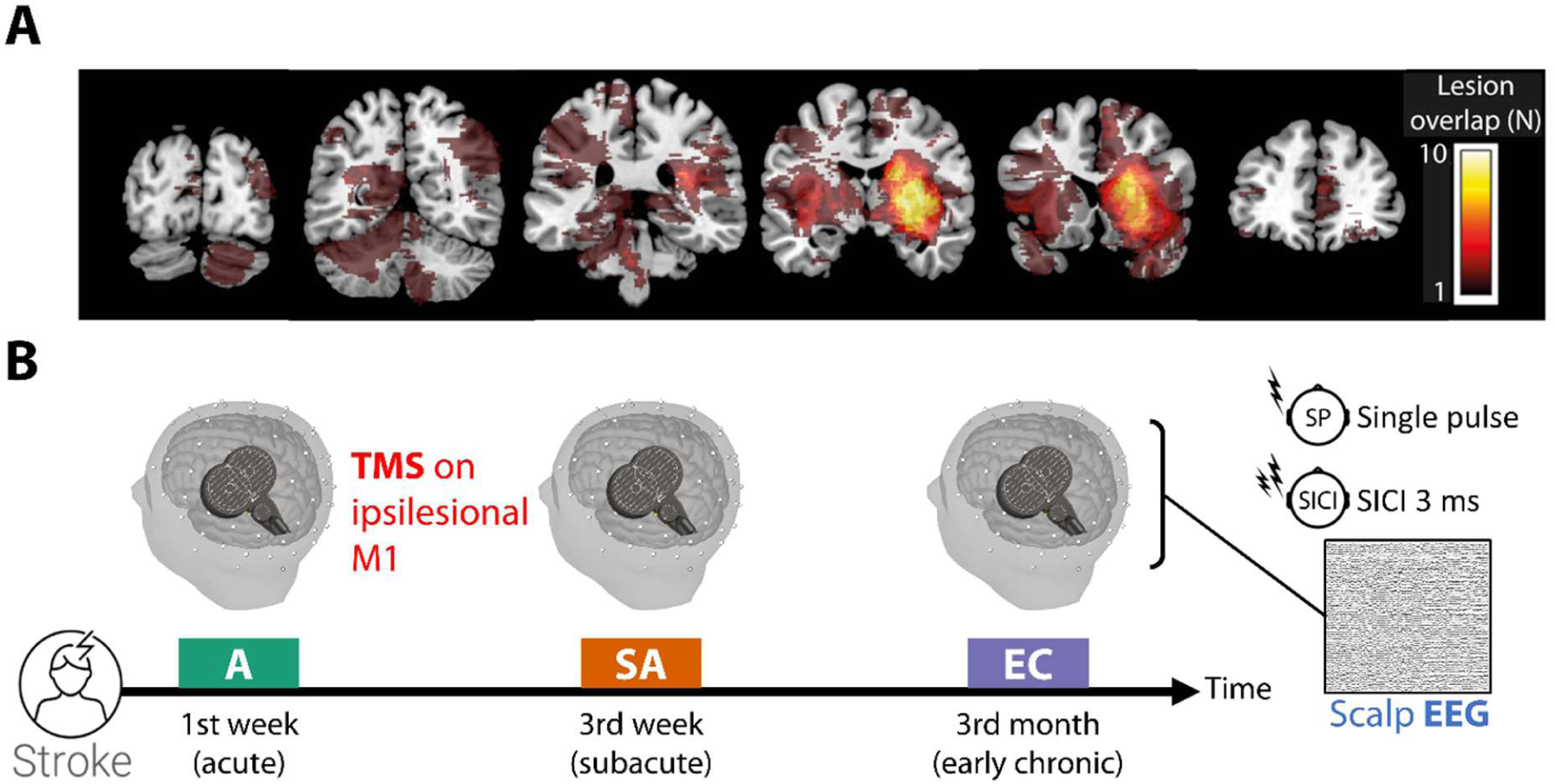
Lesion heat map and protocol design. **A.** Lesion heat map of the patients in the acute stage, N = 54. Please note that several patients of the cohort did not undergo MRI in the acute stage. **B**. Protocol design. Patients underwent 3 TMS-EEG recording sessions in the acute (A, 1^st^ week post-stroke), subacute (SA, 3^rd^ week post-stroke) and early chronic stages (EC, 3^rd^ month post-stroke). EEG was recorded while stimulating the ipsilesional motor cortex using supratreshold single pulses (SP condition) and paired-pulse short-interval intracortical inhibition protocol (SICI condition) using a 3 ms inter-stimulus interval.

**Table 1.** Patients’ characteristics. MEP, rMT and FM-UL stand for Motor evoked potential, resting Motor Treshold and Fugl-Meyer of the upper limb, respectively.

| Gender | Handedness | Stroke type | Hemisphere affected | MEP negative at T1 | rMT A (% MSO) | FM UL A (/60) | FM UL SA (/60) | FM UL EC (/60) | Days post-stroke A | Days post-stroke SA | Days post-stroke EC |
| --- | --- | --- | --- | --- | --- | --- | --- | --- | --- | --- | --- |
| 18 F / 48 M | 55 right-handed / 8 ambidextrous / 3 left-handed | 63 ischemic / 3 hemorrhagic; 57 first-ever / 9 recurrent | 32 Left / 34 Right | N = 8 | 43 ± 9.8 | 46.8 ± 19 | 50.6 ± 17 | 55.1 ± 12 | 6.6 ± 2.3 | 26.9 ± 4.9 | 98.1 ± 8.6 |

### Protocol design

Patients underwent three sessions of assessments, at one-week (acute stage: A), three weeks (subacute stage: SA) and three months (early chronic stage: EC) post-stroke (Fig. 1.B). Each session comprised structural and functional MRI, resting-state EEG and TMS-EEG, as well as a comprehensive battery of cognitive and motor evaluations. For more details on the protocol and analysis, the reader might refer to Fleury *et al.*^37^.

### Behavioral assessment

The behavioral evaluation battery comprised of the (i) Fugl-Meyer of the upper limb (FM-UL, max 60 points without reflexes) and each of its subscores: the upper extremity (FM-UE, max 32 points), the hand and the wrist^39^. For each hand, the following was assessed: (ii) the maximum fist, key and pinch force assessed in three trials and performed using a JAMAR® hydraulic hand dynamometer^40^; (iii) the Box and Blocks^41^; (iv) the nine-hole peg ^42^ (9HP). For every motor score, with the exception of the FM, a ratio between the performance of the affected and non-affected hand (affected/unaffected) was used for the analyses.

### Experimental procedure

TMS-EEG data acquisition is detailed in the third data sheet of Fleury *et al.*^37^. All the recommendations from international guidelines on TMS-EEG acquisition were followed, with the exception of a sham stimulation^43^. Two types of stimulation were applied on the ipsilesional motor cortex: single-pulse (SP) and SICI, comprised of a conditioning pulse at 80 % of the resting Motor Threshold followed by a suprathreshold SP (intensity corresponding to 1 mV MEP), with an inter-stimulus interval of 3 ms. For each patient and stroke stage, maximum of 180 SP and 180 SICI trials were applied in six separated blocks (mean 169, minimum 80). During each block, the order between conditions (SP or SICI) was pseudo-randomized.

### Data analysis

EEG data were analyzed on Matlab (MathWorks, Massachusetts, USA) with EEGLAB^44^ and the TESA toolbox^45^. TMS-EEG data preprocessing is detailed in Fleury *et al.*^37^. The mean number of ICA components removed was 8.5. For the purpose of visualization and cluster-based permutation statistics (see *Statistics*), topographies were flipped as necessary, ensuring that the left hemisphere was designated as the ipsilesional hemisphere for all patients. Local mean field power (LMFP)^46^ was computed based on the five electrodes closest to the stimulation target (FC3-FC1-C3-C1-Cz or FC4-FC2-C4-C2-Cz depending on the lesioned hemisphere). To quantify early activity power, the LMFP was summed from +20 to +80 ms after TMS. Regression quality scores (RQS) were computed to assess changes in response dynamics^19,47^. In short, the local TEPs were derived for each stroke stage within the +20 to +80 ms, and were linearly regressed on single trials of the other stroke stages. RQS is finally defined as the t-statistic associated to the local TEP factor, averaged across trials for each stroke stage and patient. RQS computed from different stroke stages allow assessing the level of similarity between the respective response dynamics. In order to quantify the complexity of the signal, we measured the number of deflections (N_def._) in the first 200 ms of the local TEP from the electrode C3 or C4^29^. For that purpose, an automated detection of significant peaks was used. Using the *find_peaks* function from the *scipy* python toolbox, we calculated the average peak-to-peak amplitude from every detected peak in the 200 ms baseline before the TMS pulse. Any peak post-TMS higher than 2 standard deviations from this average was considered as significant.

### Statistics

The early part of the TEPs from healthy older adults and patients in the acute stage were compared using cluster-based permutations, run on the +20 to +100 ms post-TMS period (using two-tailed independent t-test and a significance threshold of α = 0.05 at the sample level, and a minimum of 2 neighboring channels and 1,000 permutations at the cluster level). The significance threshold for clusters was set to p_cluster_ < 0.05. All remaining statistical analyses were performed using the JASP software (JASP Team (2022), Version 0.17.2.1). Bayes factors (BF_10_) were used to quantify statistical evidence^48^, and default values for priors were kept. LMFP was evaluated using Bayesian one-way ANOVA, for comparing patients to healthy older adults, and ANCOVA. This latter was focused on patients and comprised the stroke stages as fixed factors, the patients as random factors and the FM-UL and the supra-threshold TMS intensity as covariates. RQS was analyzed using two-way ANOVA, with strokes stages corresponding to reference TEPs and trials taken as factors. Additionally, at each stroke stage we performed Bayesian correlations between each pair of TMS-EEG readouts and behavioral scores. As the distributions of the motor scores in our patient cohort were not normal, Kendall’s nonparametric correlations were performed, and the 95% credible interval for τ_b_ (better adjusted for ties than τ_a_) were reported, referred to thereafter as τ. Evolution between stroke stages was measured as a percentage of change (e.g., A vs. SA: (𝑥𝑆𝐴 − 𝑥𝐴)/𝑥𝐴) for each behavioral score and electrophysiological readout. In order to evaluate the influence of the conditioning pulse in the SICI paradigm over LMFP, we calculated the arithmetic difference between SP and SICI (LMFP SP – LMFP SICI).

### Voxel lesion mapping

Voxel lesion symptom mapping (VLSM) was performed in order to investigate the relationship between the different TMS-EEG readouts and lesion sites. This analysis was conducted using T1 MPRAGE lesion maps with the NiiStat toolbox. The number of permutations was fixed to 2,000. Only voxels presented in at least 10% of the patients were considered. The analysis was restricted to regions of interest from the AAL^49^ and CAT^50^ atlases, in order to include both cortical and subcortical structures.

## Results

Overall, TMS-EEG recordings were performed in 60 acute, 48 subacute and 37 early chronic patients. Not all patients completed every session due to time constraints with clinical evaluations, patient unavailability, COVID-19 pandemic or the introduction of an exclusion factor, such as benzodiazepine intake. All the patients went through the protocol without reporting any adverse effects.

### Brain reactivity of acute stroke patients

Acute stroke patients presented an abnormal brain reactivity when probing the ipsilesional motor cortex, compared with the healthy older adults. Grand average TEPs revealed a larger and simpler pattern within the early part of the response (< 100 ms) in the patient cohort (Fig. 2.A). This difference was significant at the group level, as shown by cluster-based permutation statistics, for both SP (p_cluster_ < 0.05, t = +31 to +100 ms) and SICI conditions (p_cluster_ < 0.05, t = +30 to +57 ms; Fig. 2.A). When exploring the data at the individual level, this abnormal reactivity seemed to be more pronounced in severely affected patients (Fig. 2.B). This inter-patient variability was also evident when investigating the local response of the ipsilesional motor cortex longitudinally (Fig. 3.A & 4.A). As a means to assess (i) the cortical excitability and (ii) the complexity of the TMS-evoked dynamics, we computed for each stroke stage (i) the LMFP of the early response (< 80 ms), and (ii) the number of evoked deflections and the RQS between stroke stages.

**Figure 2.**
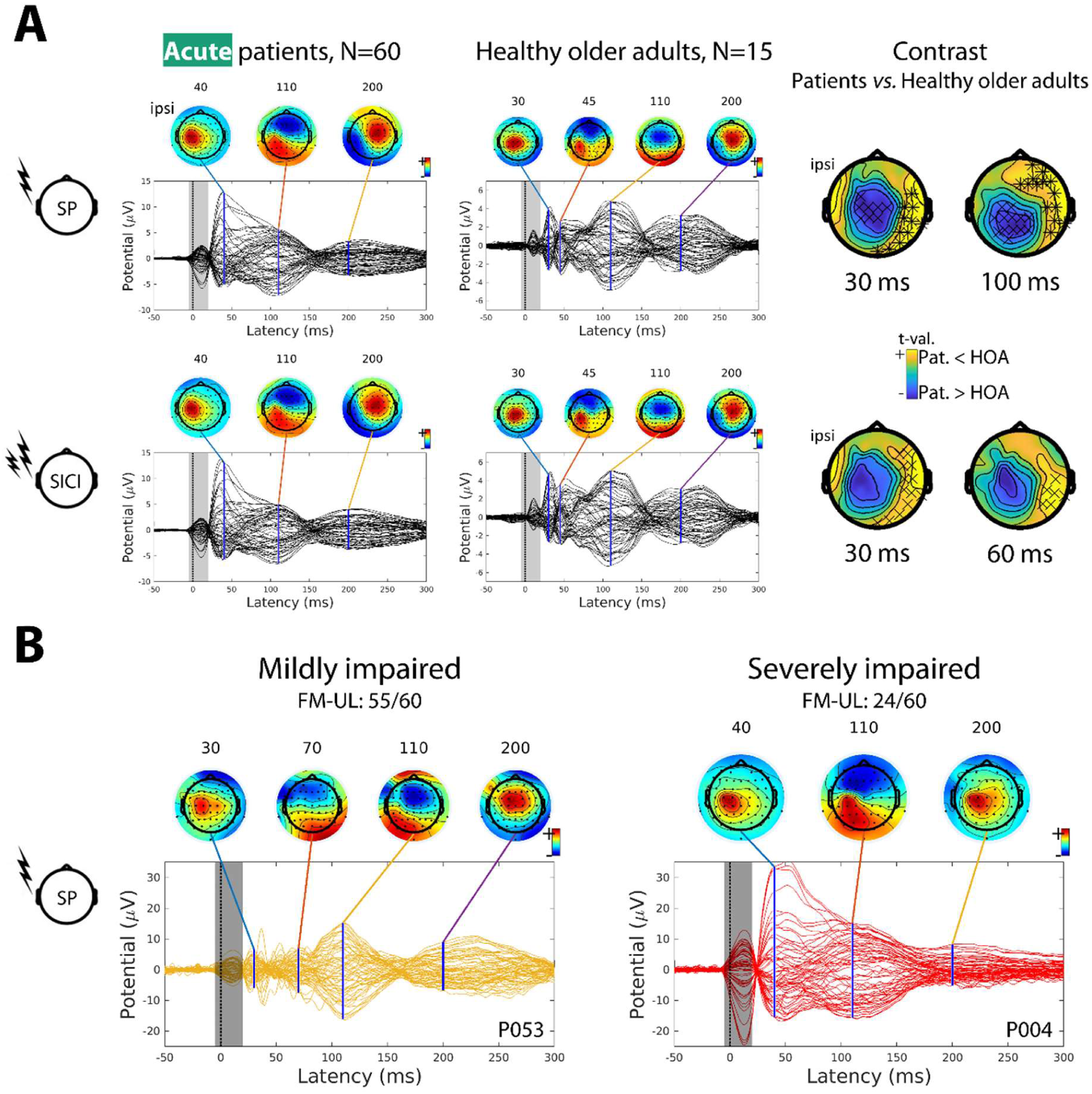
Abnormal brain reactivity in acute stroke patients after the stimulation of the ipsilesional motor cortex. **A.** Grand average TMS-evoked potentials (TEPs) of the patient cohort in the acute stage (left column), and of the healthy older adults (right column), for each stimulation condition, i.e., suprathreshold single pulse (SP, top row) and short-interval intracortical inhibition (SICI, bottom row). Left hemisphere was designated as the ipsilesional hemisphere for all patients (see text). Electrodes’ timeseries are overlayed in a butterfly view from −50 to +300 ms relative to the stimulation onset, and topographies are plotted for the main activity peaks, the colormap being adjusted to each temporal min and max value. The grey shaded area represents the time window interpolated around the TMS pulse (from −5 to + 20 ms) not taken in the analysis. Topographies in the right column depict the beginning (left) and end (right) of the significant clusters found when contrasting TEPs between patients and healthy controls within the first 100 ms. Colormap codes for the local t-value, and black crosses and stars indicate electrodes belonging to a significant cluster (as measured by cluster-based permutation tests, see *Statistics* section). **B.** Examples of TEPs in two representative patients in the acute stage with different initial motor deficit, as indicated by the Fugl-Meyer score of the affected upper limb (FM-UL). Comparing TEPs reveals the difference in both maximum amplitude and spatial distribution of the signals, with the most affected patient (right) exhibiting a simpler, larger and more spatially restricted activity, especially during the first 100 ms.

**Figure 3.**
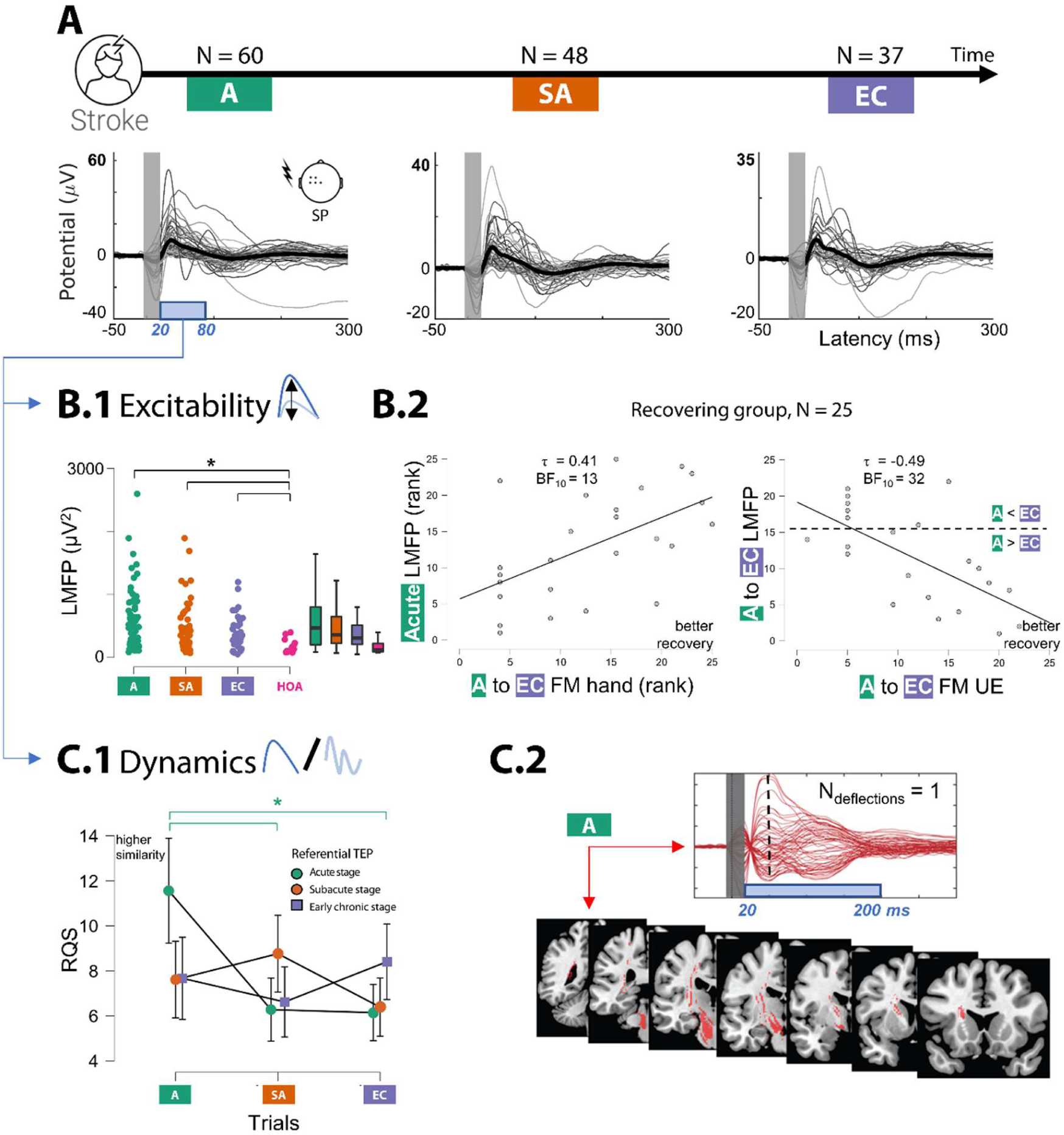
Longitudinal evolution of motor cortical excitability and TMS-evoked dynamics, and their association with motor recovery. **A.** Local TEPs from the ipsilesional electrodes close to the stimulation site, across the three timepoints, for each patient (one line represents one patient). Please note the decrease of max amplitude through stroke stages, and the inter-patients’ variability in the evoked response amplitude and dynamics, respectively assessed using LMFP and RQS on the early part of the response (20-80 ms). **B.1.** Distribution of LMFP across stroke stages, each being significantly higher than the one from healthy older adults (HOA), as indicated by asterisks (see text). **B.2.** Significant associations between high LMFP in the acute stage (left), and strong LMFP decrease towards the early chronic stage (right) on one hand, and better motor recovery on the other hand in the recovering group. Such correlations were also found within the whole cohort of patients, with weaker level of statistical evidence (see Table 2). Correlations were performed using Kendall’s τ_b_, which are better adjusted for ties; values displayed on both axes corresponding to ranks. **C.1.** Distribution of RQS through stroke stages (y axis), when using the local TEP of each stroke stage as a reference (represented by a different line). Higher RQS highlight higher similarity between the evoked dynamics of the reference TEP and the trials from other stroke stages. Stars indicate significant post-hoc tests revealing strong evidence for a difference in evoked dynamics between the acute and both subacute and early chronic stages. **C.2.** Association between response features and lesions maps were assessed using a Voxel Lesion Symptoms Mapping (VLSM). In the acute stage, the number of deflections within the first 200 ms was found negatively correlated with lesions in the internal capsule (depicted by red voxels).

**Figure 4.**
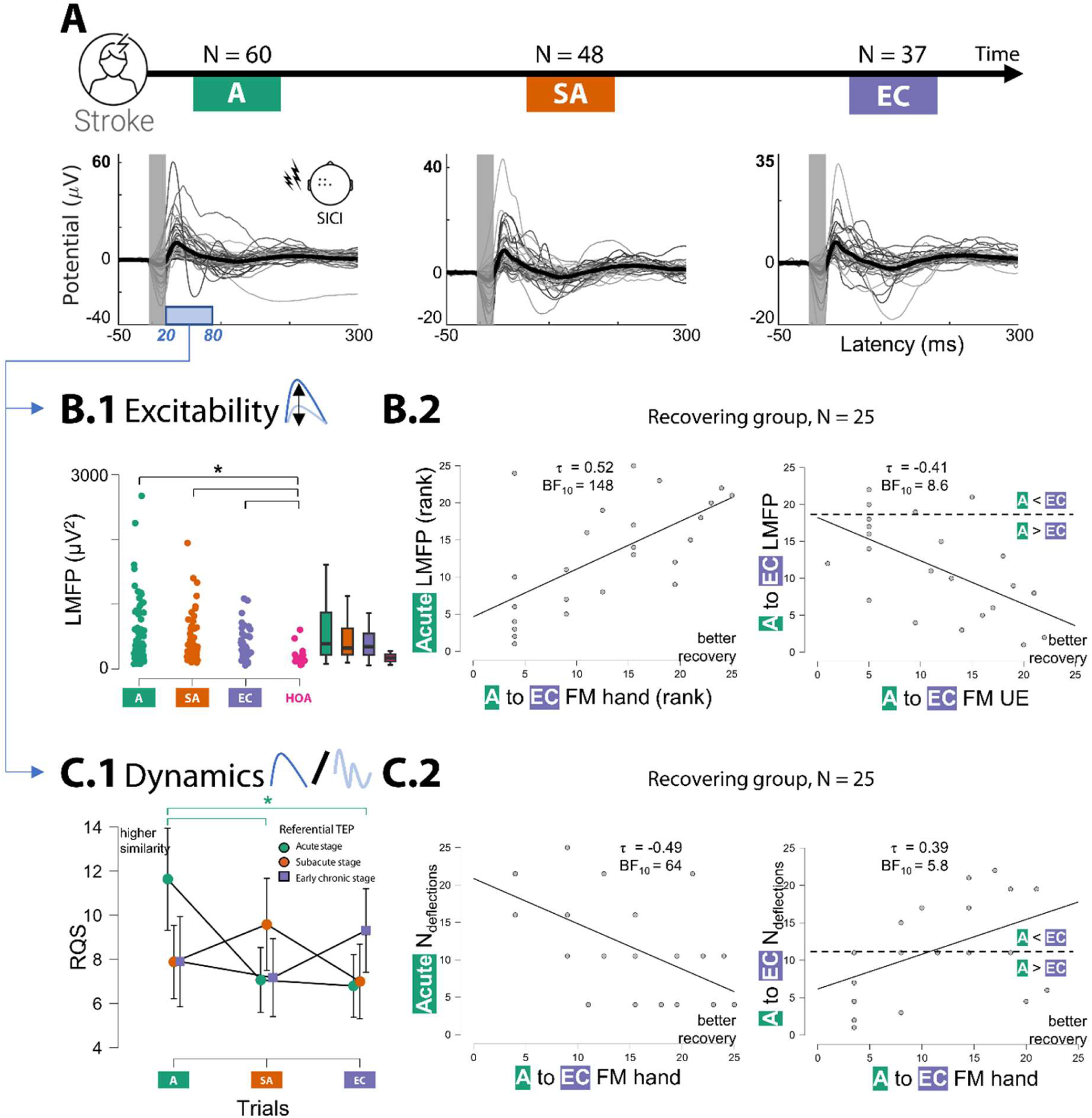
Longitudinal evolution of motor intracortical inhibitory circuits excitability and dynamics, and their association with motor recovery. **A.** Local SICI-evoked potentials from the ipsilesional electrodes close to the stimulation site, across the three stroke stages, for each patient (one line represents one patient). Please note the decrease of max amplitude through stroke stages, and the inter-patients’ variability in the evoked response amplitude and dynamics assessed using LMFP and RQS respectively, on the early part of the response (20-80 ms). **B.1.** Distribution of SICI-evoked LMFP across stroke stages, each being significantly higher than the one from the healthy older adults (HOA), as indicated by asterisks (see text). **B.2.** Significant associations between high SICI-evoked LMFP in the acute stage (left), and strong SICI-evoked LMFP decrease towards the early chronic stages (right) on the one hand, and better motor recovery on the other hand in the recovering group. Such correlations were also found within the whole cohort of patients, with weaker level of statistical evidence (see Table 2). Correlations were performed using Kendall’s τ, values displayed on both axes corresponding to ranks. **C.1.** Distribution of RQS through stroke stages (y axis), when using the local TEP of each stroke stage as a reference (represented by a different line). Higher RQS highlight higher similarity between the SICI-evoked dynamics of the reference TEP and the trials from other stroke stages. Stars indicate significant post-hoc tests revealing strong evidence for a difference in SICI-evoked dynamics between the acute and both subacute and early chronic stages. **C.2.** Significant associations between SICI-evoked dynamics, as assessed by the number of deflections within the first 200 ms, and motor recovery. A low number of deflections in the acute stage (left), and an increase of deflections towards the early chronic stage (right), were associated with positive motor outcomes in the early chronic stage. Such correlations were also found within the whole cohort of patients, with stronger level of statistical evidence (see Table 2).

**Table 2.**
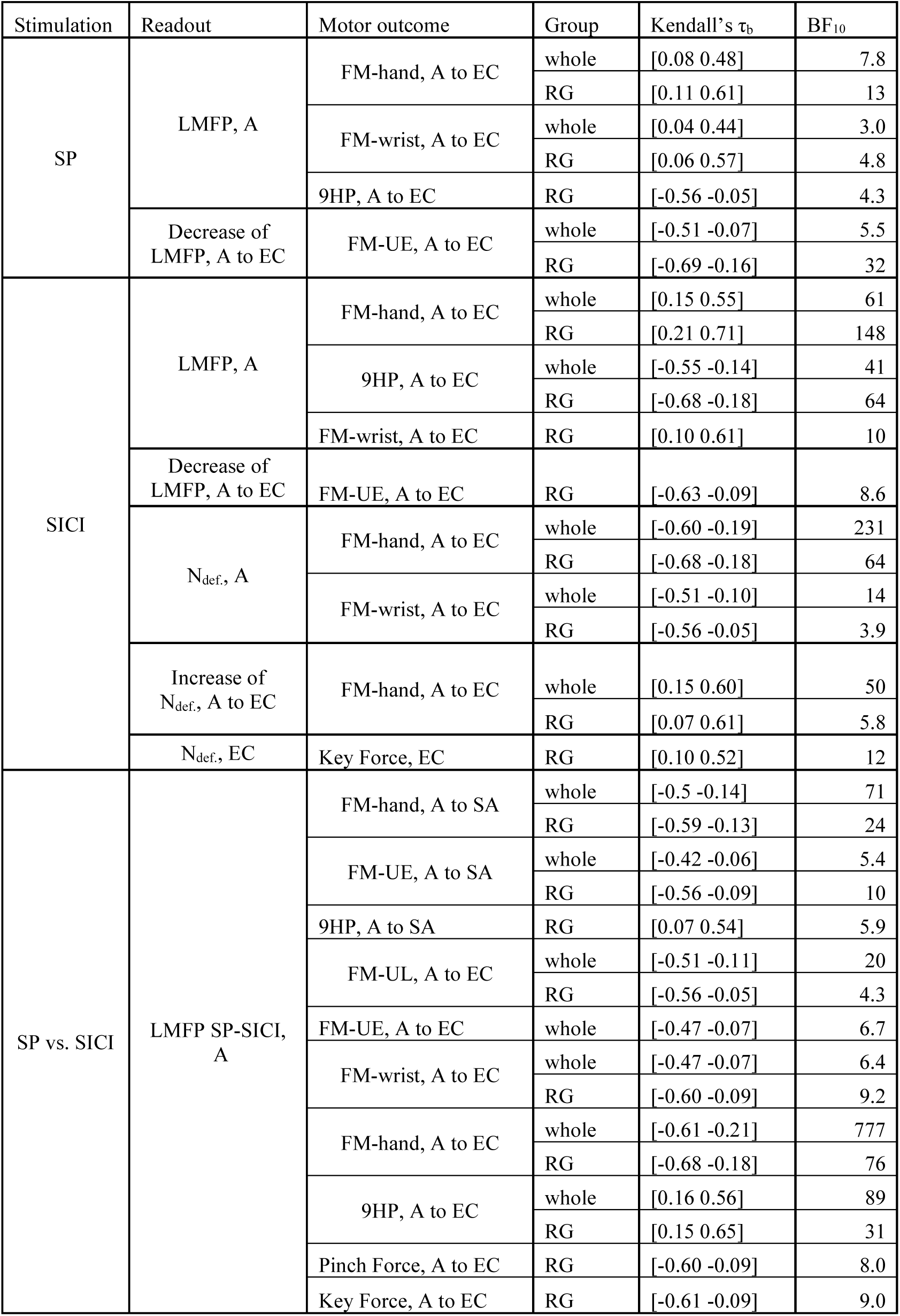
Correlations between TMS-EEG readouts and motor outcomes. *Whole* and *RG* flag correlations found within the whole cohort of patients and the recovering group respectively. *N_def._* stands for number of signal’s deflections (see *Data analysis* section).

### Motor cortical excitability across stroke stages

At the group level, the LMFP of the early response was higher in stroke patients than healthy controls for each stroke stage (Fig. 3.B.1), as shown with moderate evidence by the Bayesian one-way ANOVA (BF_incl_ = 7.0 for the group effect) and with moderate to strong evidence by the subsequent post-hoc comparisons (BF_10_ = 13, 12, and 9.4 when comparing controls to acute, subacute, and early chronic stages respectively). However, despite a visual trend of decreased LMFP with time, post-hoc comparison did not reveal any significant difference between stroke stages (0.1 < all BF_10_ < 1). This was further confirmed by the Bayesian ANCOVA focused on stroke patients: the level of statistical evidence was inconclusive regarding stroke stages at the group level (BF_10_ = 0.7). However, it revealed strong evidence for a link between the LMFP and its covariate FM-UL (BF_10_ = 16; using FM-UE: BF_10_ = 33), with larger signal power being associated with reduced upper limb scores (CI_95%_: [−14.7, −5.2]). There was also extreme evidence for an effect of the TMS intensity used (BF_10_ > 1.4.10^4^), with higher LMFP linked with higher intensity (CI_95%_: [6.6, 19.1]). Moreover, abnormally high LMFP in the acute stage and its decrease with time were positively correlated with motor recovery towards the early chronic stage (Fig. 3.B.2., Table 2).

### TMS-evoked dynamics of the motor cortex across stroke stages

The study of TMS-evoked dynamics revealed that their characteristics were singular in the acute stage. First, ANOVA investigating the evoked dynamics in different stroke stages, captured by RQS, revealed extreme evidence for an interaction effect between the factors reference TEP and single trial activity (BF_10_ > 1.10^8^, Fig. 3.C.1). Post-hoc tests showed strong evidence for a difference between stroke stages only when using the TEP from the acute stage as a reference (A > SA, BF_10_ = 44; A > EC, BF_10_ = 23). No evidence was found when using TEPs from the other stroke stages (all BF_10_ ∈ [0.2 1.3]). Regarding the number of deflections, no effect was found for the factors stroke stages, FM-UL score or TMS intensity. Similarly, no evidence of relationships between the number of deflections and any of the motor scores was found. VLSM with the number of deflections in the acute stage revealed an association with the internal capsule (Fig. 3.C.2). Thus, simpler responses, i.e., fewer deflections, were predominantly related with lesions in the corticospinal tract. There was no evidence for presence or absence of any correlations between these readouts and motor scores.

### Motor intracortical inhibitory circuits excitability and dynamics

Overall, using the same analysis framework as for SP, probing the GABAergic system with SICI did not modulate the previous readouts. Stroke patients still exhibited higher LMFP compared with healthy controls in each stroke stage (Fig. 4.B.1). This was supported by lower moderate evidence from Bayesian ANOVA (BF_incl_ = 3.2) and from post-hoc comparisons (BF10 = 5.8, 4.3, and 4.9 when comparing controls to acute, subacute, and early chronic stages, respectively). Bayesian ANCOVA focused on stroke patients further confirmed the lack of conclusive evidence regarding stroke stages (BF_10_ = 1.2), but revealed very strong evidence for an effect of the FM-UL (BF_10_ = 44; using FM-UE: BF_10_ = 176) and of the TMS intensity (BF_10_ = 598). The Bayesian correlations also showed links between acute higher LMFP in SICI and its decrease over time on the one hand, and greater motor recovery on the other hand (Fig. 4.B.2, Table 2). No effect was found when contrasting LMFP from SP and SICI. Regarding TMS-evoked dynamics, the RQS analysis also demonstrated an atypical dynamical signature in the acute stage compared with the following timepoints (interaction effect: BF_10_ > 6.10^7^; all post-hoc BF_10_ ∈ [9 12] when taking acute TEP as reference, BF_10_ ∈ [0.2 0.9] otherwise, Fig. 4.C.1). Second, patients who recovered more exhibited fewer deflections in the acute stage and a greater increase in the complexity of the evoked signal along time post stroke (Fig. 4.C.2, Table 2).

### Unmasked complementary intracortical inhibition mechanisms

The association between the intracortical inhibition effect on excitability, i.e., by contrasting LMFP from SP and SICI conditions, and motor improvement was only found when removing the abnormal large component (Fig. 5.A). This component was detected in 34/60 (57%), 27/48 (56%), 17/37 (46%) in the acute, subacute, and early chronic stages respectively. First, moderate evidence was found for an absence of difference between healthy older adults and patients in any stroke stages (BF_incl_ = 0.2, Fig. 5.B.1). Second, the Bayesian ANCOVA focused on patients failed to find any evidence for an effect of stroke stage, FM-UL or TMS intensity (0.26 < all BF_incl_ < 0.89). However, moderate to very strong evidence was found for a negative link between the contrast in LMFP in the acute stage and motor recovery: patients who recovered the most were those presenting disinhibition, i.e., higher level of LMFP in SICI than in SP (Fig. 5.B.2, Table 2). Longitudinal changes within the dynamics of the SICI-evoked response were also revealed in the recovering group. When comparing the dynamics in the acute and subacute stages using RQS, moderate evidence was found with long-term motor improvement in the early chronic stage (FM hand, τ ∈ [−0.66 −0.07], BF_10_ = 5.8; 9HP, τ ∈ [0.09 0.67], BF_10_ = 7.3) (Fig. 5.C). Stronger motor improvements between the acute and the following stroke stages were linked with greater differences in the pattern of the SICI-evoked activity along time.

**Figure 5.**
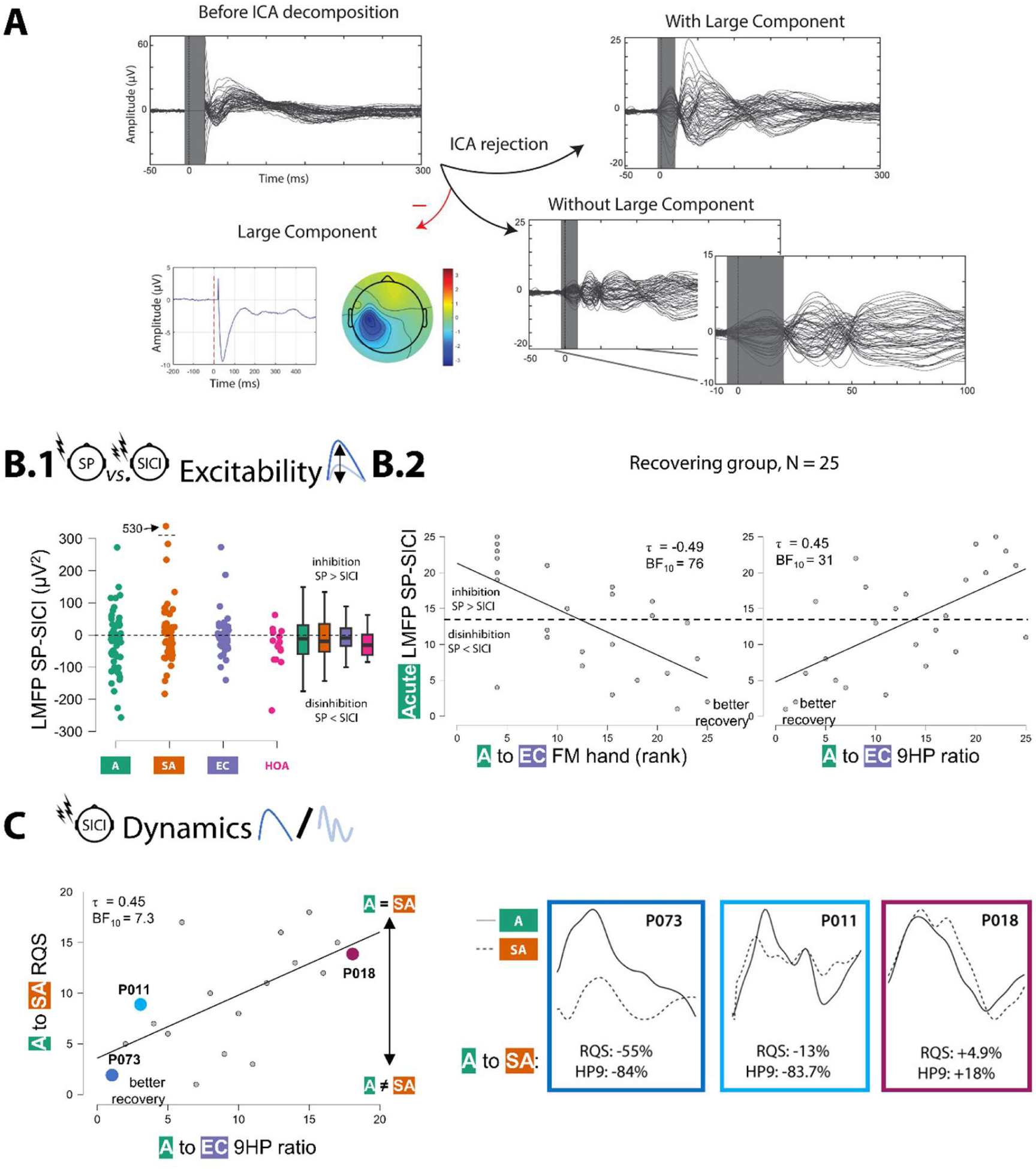
Complementary intracortical inhibition mechanisms revealed after removal of the abnormal large component. **A.** The large and monophasic evoked component, when visually detected, was removed during the second round of ICA decomposition prior to these complementary analyses. The removal of the large component (bottom left) unmasked weaker and more complex evoked signals (bottom right, “without large component”). **B.1.** Distribution of the contrast between LMFP of SP and SICI protocol across stroke stages, and for healthy older adults (HOA). On average, values were negative, i.e., early LMFP was higher after SICI than SP, indicating a tendency towards disinhibition^19,51–54^. **B.2.** Significant associations between high disinhibition in the acute stage (left), and better motor recovery in the recovering group. Such correlations were also found within the whole cohort of patients, with stronger level of statistical evidence (see Table 2). Correlations were performed using Kendall’s τ, values displayed on both axes corresponding to ranks. **C.** Significant association between change in RQS scores for SICI condition from the acute to the subacute stages (y axis) and motor recovery toward the early chronic stage (left). Plots of individual local TEPs after SICI stimulation in the acute (solid line) and subacute (dotted line) stages for three representative patients. Respective evolution in RQS and 9HP (a decrease means improvement) are expressed below each signal. Note that a low value of RQS change between stroke stages correspond to a high similarity in the evoked signals dynamics, i.e., no evolution in evoked dynamics, between these stages, which is associated with worse recovery (P018, right).

## Discussion

### Abnormal brain reactivity in stroke patients

Our results suggest that stroke patients present atypical brain reactivity patterns, when the ipsilesional motor cortex is stimulated by means of TMS. In the acute stage, the pathological response was characterized by a large monophasic response, which contrasted with the weaker and more complex response observed in healthy young^47^ and older adults (Fig. 2). This result extends previous findings that spotted this abnormal reactivity in severely affected stroke patients^29,55^, other cortical injuries^30^, and altered brain states^56^. Results centred on early LMFP indicated that stroke presented an hyperexcitable ipsilesional motor cortex compared with age-matched controls, or with what is generally found in healthy young^18^ and older^32,57^ populations (Fig. 3.B.1). Interestingly, the high inter-patient variability regarding excitability explained the level of motor impairment: this hyperexcitability was mostly found in the most affected patients and was positively linked with the level of motor impairment (Fig. 3.B.2).

Analysis of the evoked response’s dynamics indicated that part of this abnormal reactivity was specific to the acute stage and evolved through stroke stages (Fig. 3.C.1). Signal dynamics being sensitive to both local, e.g., cytoarchitectonics and neurotransmitter concentrations^22,26^, and global properties, e.g., structural and functional connectivity^23^, such results could highlight the post-stroke reorganization processes occurring at these various levels. Regarding the global level, the VLSM revealed that acute signal dynamics were associated with the lesion load in the internal capsule (Fig. 3.C.2), hosting the main outflow from motor cortical areas containing fibers from the corticospinal, corticorubral and corticopontine tracts^50^, in line with previous work^29^. Such disruption of fibers connecting the cortex to subcortical structures prevents propagation and integration of the evoked activity to distant brain areas, leading to simpler responses reflected by fewer deflections. Recent work has succeeded this effect, showing that lesioned structural connectomes tended to produce simple and local TMS-evoked responses^58^. However, the longitudinal changes observed thereafter might rather underline reorganization processes occurring at the level of local intracortical inhibitory systems.

### Disinhibition as a key mechanism for successful recovery

Recent animal work suggested that hyperexcitability and disinhibition states occur between the first week and one-month post-stroke and plays an essential role for neuronal plasticity and recovery^59–61^. Indeed, in the acute stage, GABA-mediated ipsilesional intracortical inhibition is reduced compared with the unaffected hemisphere and to healthy controls^62^. Animal models^6,63,64^ and human studies^65–67^ suggest that this acute disinhibition is adaptive by enhancing ipsilesional neuronal excitability through reduction of cortical inhibition. This decrease of cortical inhibition is thought to promote plastic changes and reorganization to sustain recovery of the lost functions^17,68^. If indirect proof of such disinhibition lies in the abnormally strong response observed here, the use of the SICI stimulation protocol enabled the direct probing of the intracortical GABAergic inhibitory system. In healthy young adults, SICI is known to induce an inhibition of the early TMS-evoked cortical activity^19,51–54^. In opposite, our results showed that SICI-induced inhibition was perturbated in the patient cohort, which showed strong inter-individual variability with a tendency toward disinhibition at the group level (Fig. 5.B.1).

The adaptive or maladaptive nature of this process remains largely unknown. In the present study, the acute disinhibition was associated with better recovery of distal impairment and fine motor skills, especially in the recovering group (Fig. 5.B.2). These findings point to the fact that fine-tuned inhibitory activity is especially critical for more skilled hand functions, e.g., represented by the 9HP test. Although Tscherpel *et al.*^29^ also showed a relation between a large and simple reactivity in the acute stage and motor recovery, the direction of the effect was opposite to the present results. However, the link between abnormal reactivity and worse improvement reported in the previous study could be explained by a greater proportion of severely affected patients with limited improvement (7 out of 25 remained at 0 in ARAT at 3-months post stroke). While the present cohort contained a higher proportion of mildly to moderately affected patients, with only 2 out of 66 having 0 in ARAT in the early chronic stage, it is possible that the mildly impaired patients exhibited the same physiological response to TMS, but showed more recovery due to their overall less impaired initial status. Finally, a disinhibition state was also present in our healthy older adults’ group, which did not significantly differ from the patients (Fig. 5.B.1). The perturbation of the GABAergic inhibitory system with age is known^57,69,70^, and has been linked to the deficit in inhibitory control^71^. Further investigation would be needed to determine whether the acute beneficial disinhibition observed here is a direct result of post-stroke mechanisms, or whether it highlights the anterior presence of a “natural” age-induced disinhibition that proved beneficial after stroke.

Interestingly, such beneficial disinhibition was also found in the present cohort of patients when focusing on late TMS-induced alpha oscillations^38^, which is also a marker of GABAergic system activity^72^. The time course of this disinhibition, i.e., between the subacute and early chronic stages, and spatial localization, i.e., more diffused across the brain, differed from the acute and local phenomenon found here. Taken all together, our results suggest that motor recovery is supported by a disinhibition phase occurring first in the acute stage to promote plastic changes and reorganization of the localized impacted areas, which then spread on a larger scale to allow remote plasticity and network reorganization towards the early chronic stage^8,73,74^.

### Changes of intracortical inhibitory activity within ipsilesional motor cortex and motor recovery

Previous work has hypothesized different roles for persistent disinhibition in the chronic stage. While Ding *et al*.^75^ speculated that disinhibition could be detrimental for motor recovery, other studies showed that a persistent disinhibition in the chronic stage might support recovery through enhanced plasticity in patients with residual deficits^17,76,77^. The functional role of persistent disinhibition in the chronic stage is thus unclear and the present longitudinal data contributed to addressing this question. Several readouts showed an evolution associated with motor recovery and our results rather pointed toward a beneficial decrease of the disinhibition, i.e., a restoring of a typical intracortical inhibitory activity within the ipsilesional motor cortex.

First, the association between the response power decrease and motor recovery constitutes indirect evidence for a restoration of local inhibitory activity reducing cortical excitability (Fig. 3.B.2). More direct evidence was found from the analysis of the SICI-evoked dynamics. First, the rapid change in evoked dynamics between the acute and subacute stages was correlated with future motor recovery (Fig. 5.C). Second, the number of signal deflections increased with respect to motor recovery towards the early chronic stage, indicating a return to more complex response patterns for the recovering patients (Fig. 4.C.2). Such changes in signal dynamics, that were absent when the motor cortex was probed using single pulses, might highlight the functional reorganization that is at stake within the ipsilesional and intracortical GABAergic inhibitory system. The influence of larger scale structural and functional reorganization occurring post-stroke^78^ cannot however be ruled out. The changes in dynamics observed here, and especially the increase in the number of deflections, might reveal the re-emergence of recurrent large-scale networks^58^, that might be specifically linked to inhibitory functions (e.g., inhibitory control network^79^).

### Unmasking complementary intracortical inhibition mechanisms by removing atypical large evoked activity

We hypothesized that the large observed responses might mask further underlying intracortical inhibition mechanisms supported by neuronal activity of weaker power. In fact, the results related to excitability and early dynamics readouts were of the same nature for SICI as for single pulse (Fig. 3 & 4). To do so, we removed the corresponding large component during the second round of ICA (Fig. 5.A), which unveiled complementary information on the intracortical inhibitory system. We propose that such large activity might predominantly originate from large layer V (L5) pyramidal cells present in the motor cortex, since both recording and stimulation techniques used here are biased toward this neuronal population^80–82^. On the one hand, these cells are known to generate the strongest electrical activity, at both microscale and mesoscale^81,83^, while being the most excitable neurons^84^. On the other hand, their specific shape and spatial orientation within the gyrus makes them more sensitive to the electrical field induced by TMS^80,82^. Therefore, TMS-EEG coupling might be oversensitive to deep L5 pyramidal neurons population and removing this large activity might allow to reveal activity from neurons within superficial layers eliciting weaker electrical potentials, such as inhibitory interneurons.

### Limitations

Although this study followed most recent guidelines on TMS-EEG coupling acquisition and data processing^43^, the absence of a realistic sham stimulation condition prevented the assessment of the influence of peripheral evoked potentials, resulting from the multisensory nature of TMS^85,86^. However, since this study mostly focused on early components, which are known to be much less sensitive to peripheral influence^19,47,87,88^, and employed a longitudinal analysis approach with the comparison with a proper age-matched control group, the absence of a sham stimulation condition does not affect the final interpretation of the results. Another limitation pertains to the distribution of the patient cohort concerning the severity of motor impairment in the acute stage and the extent of recovery. The majority of patients exhibited moderate to mild impairment. Conducting additional analyses on more heterogeneous groups in future studies will help refine the current conclusions regarding the relationship between changes in brain responsivity and motor recovery.

## Conclusion

In conclusion, this work offers new insights into the longitudinal changes of cortical reactivity and local intracortical inhibition in the affected motor cortex after a stroke. The present results strongly support the critical impact of intracortical disinhibition evolving in the acute stage on residual motor function and recovery, especially skilled distal functions, and the importance of reorganization within the intracortical inhibition system of the lesioned motor cortex to sustain long-term motor recovery. Furthermore, this study demonstrates that TMS-EEG provides an excellent opportunity to determine the reactivity of the affected motor cortex after a stroke, even in patients with impacted corticospinal tract preventing the monitoring of peripheral motor activity with EMG. This knowledge provides a strong basis for developing TMS-EEG towards a clinical tool to phenotype patients and to develop biomarkers related to recovery and treatment response.

## Acknowledgements

This work was supported by ‘Personalized Health and Related Technologies (PHRT-#2017-205)’ of the ETH Domain, the Defitech Foundation (Strike-the-Stroke project, Morges, Switzerland), and the Wyss Center for Bio and Neuroengineering (WP030; Geneva, Switzerland), the SNSF (NIBS-iCog, 320030L_197899 / 1).

We acknowledge access to the facilities and expertise of the Center for Biomedical Imaging, a Swiss research center of excellence and of the MRI and Neuromodulation facilities of the Human Neuroscience Platform of the Foundation Campus Biotech Geneva and access to the Neuroimaging and clinical facilities of the Hopital Valais de Sion (HVS, Sion) and the Clinique romande de réadaptation (CRR, Sion).

We thank Silvia Avanzi for her excellent support during the recruitment and data acquisition process.

## Author contributions

Conceptualization: SH, ACM, TM, DvdV, MJW, PK, and FCH

Methodology: SH, ACM, TM, MW, and FCH

Investigation: SH, ACM, TM, LF, AW, MC, PM, JB, NM, EB, JA, CJ, AM, CC, VA, PV, JAG, JLT, DSM, CB, PJK, and MJD

Visualization: SH and ACM

Funding acquisition: FCH, DvdV, OB, and SM

Project administration: FCH, TM, MJW, and PJK

Supervision: FCH, PJK, MJW, and TM

Writing – original draft: SH and ACM

Writing – review & editing: all authors

## Data availability

The data related to this article and all the analysis scripts and JASP files are available upon reasonable request to the corresponding author.

